# Risk quantification for SARS-CoV-2 infection through airborne transmission in university settings

**DOI:** 10.1101/2021.03.31.21254731

**Authors:** Mythri Ambatipudi, Paola Carrillo Gonzalez, Kazi Tasnim, Jordan T. Daigle, Taisa Kulyk, Nicholas Jeffreys, Nishant Sule, Rafael Trevino, Emily M. He, David J. Mooney, Esther Koh

**Author notes:** These authors contributed equally to the paper (Paola Carrillo Gonzalez, Kazi Tasnim). These authors contributed equally to the paper (Jordan T. Daigle, Taisa Kulyk). These authors contributed equally to the paper (Emily M. He, David J. Mooney, Esther Koh).

## Abstract

The COVID-19 pandemic has significantly impacted learning as many institutions switched to remote or hybrid instruction. An in-depth assessment of the risk of infection that takes into account environmental setting and mitigation strategies is needed to make safe and informed decisions regarding reopening university spaces. A quantitative model of infection probability that accounts for space-specific parameters is presented to enable assessment of the risk in reopening university spaces at given densities. The model uses local positivity rate, room capacity, mask filtration efficiency, air exchange rate, room volume, and time spent in the space as parameters to calculate infection probabilities in teaching spaces, dining halls, dorms, and shared bathrooms. The model readily calculates infection probabilities in various university spaces, with mask filtration efficiency and air exchange rate being among the dominant variables. When applied to university spaces, this model demonstrated that, under specific conditions that are feasible to implement, in-person classes could be held in large lecture halls with an infection risk over the semester < 1%. Meal pick-ups from dining halls and the use of shared bathrooms in residential dormitories among small groups of students could also be accomplished with low risk. The results of applying this model to spaces at Harvard University (Cambridge and Allston campuses) and Stanford University are reported. Finally, a user-friendly web application was developed using this model to calculate infection probability following input of space-specific variables. The successful development of a quantitative model and its implementation through a web application may facilitate accurate assessments of infection risk in university spaces. In light of the impact of the COVID-19 pandemic on universities, this tool could provide crucial insight to students, faculty, and university officials in making informed decisions.

## INTRODUCTION

The COVID-19 pandemic has immensely impacted academia, with many higher learning institutions switching to remote or hybrid learning models for the first time (Smalley, 2020). While these policies have reduced widespread transmission, they have adversely affected learning, admissions, enrollment, athletics, campus activities, and housing. With the shift to virtual classes and students divided across the globe, challenges have arisen for instructors and students, including decreased academic engagement, syphoned access to academic resources, inability to recreate in-person activities, and obstacles for synchronous learning. A primary question university administrators face is the possibility of resuming some form of in-person classes while keeping infection risk low. This paper details how severe acute respiratory syndrome coronavirus 2 (SARS-CoV-2) infection risk is impacted by various university settings, such as dining, residential, and teaching spaces, and presents a user-friendly web application for assessing infection risk in these spaces.

Infection by SARS-CoV-2 causes COVID-19 (Yuen et al., 2020), and the mechanism of SARS-CoV-2 transmission is impacted by viral particle movement. SARS-CoV-2 virions are approximately 0.1 μm in diameter (Bar-On et al., 2020). They are emitted as droplets and aerosols by infected individuals during turbulent respiratory events (e.g., coughing and sneezing) and while breathing and speaking (Jayaweera et al., 2020; Mittal et al., 2020). Droplets are typically larger particles (>5 µm) and rapidly fall to underlying surfaces. In contrast, aerosols (particles <5 µm) evaporate faster than they fall (Augenbraun et al., 2020; Jayaweera, et al., 2020). Consequently, aerosols containing viral particles can remain suspended for hours or days and travel farther than droplets, increasing infection risk (Augenbraun et al., 2020; van Doremalen et al., 2020). Even after an infected individual has left a room, others can inhale viral particles emitted earlier. Aerosols therefore appear to pose the greatest risk of SARS-CoV-2 transmission (Augenbraun et al., 2020; van Doremalen et al., 2020). This study focuses on SARS-CoV-2 transmission through aerosols.

Key parameters impacting airborne transmission include face mask usage and ventilation. Face mask usage is recommended to reduce SARS-CoV-2 transmission (CDC, NCIRD, 2021b), as masks filter incoming airborne particles and trap outgoing respiratory secretions (Mittal et al., 2020). There are three main classifications of protective face masks: respirators, surgical masks, and cloth face coverings (The Royal Society, 2020). Different masks of varying filtration efficacy may best suit the levels of risk for particular settings, activities, or individuals. For instance, N95 respirators, which have 95% filtration efficiency, provide greater protection than surgical and cloth masks (CDC, NIOSH, 2020). Numerous studies have analyzed mask filtration efficacies and their impact on mitigating infection risk (Fischer et al., 2020; Konda et al., 2020). Aerosolized viral particles can linger in closed spaces for extended durations, so ventilation can also decrease infection risk (CDC, NCIRD, 2021a). HVAC (Heating, Ventilation, and Air Conditioning) systems aid in circulating air and diluting viral particle concentrations (EPA, 2020). HEPA (High Efficiency Particulate Air) filters, often used in combination with ventilation, can efficiently remove particles from the air.

A straightforward risk analysis method is needed to better understand the SARS-CoV-2 infection risk in university spaces. A mathematical model exists for calculating infection probabilities (Watanabe et al., 2010) and has recently been utilized for assessing SARS-CoV-2 infection risk in laboratories and office settings (Augenbraun et al., 2020). Here, that model is adapted to calculate output parameters relevant to the re-opening and occupancy of university spaces, including teaching, dining, and residential spaces. A web application has been developed that allows users to input the necessary metrics into the model, which outputs the infection probability in a given space.

## METHODS

### General Approach for Calculations in University Spaces

Approaches for calculating infection probabilities were developed for three types of university spaces: teaching spaces, dining halls, and residential spaces. These approaches followed the steps in Figure 1.

**Figure 1:**
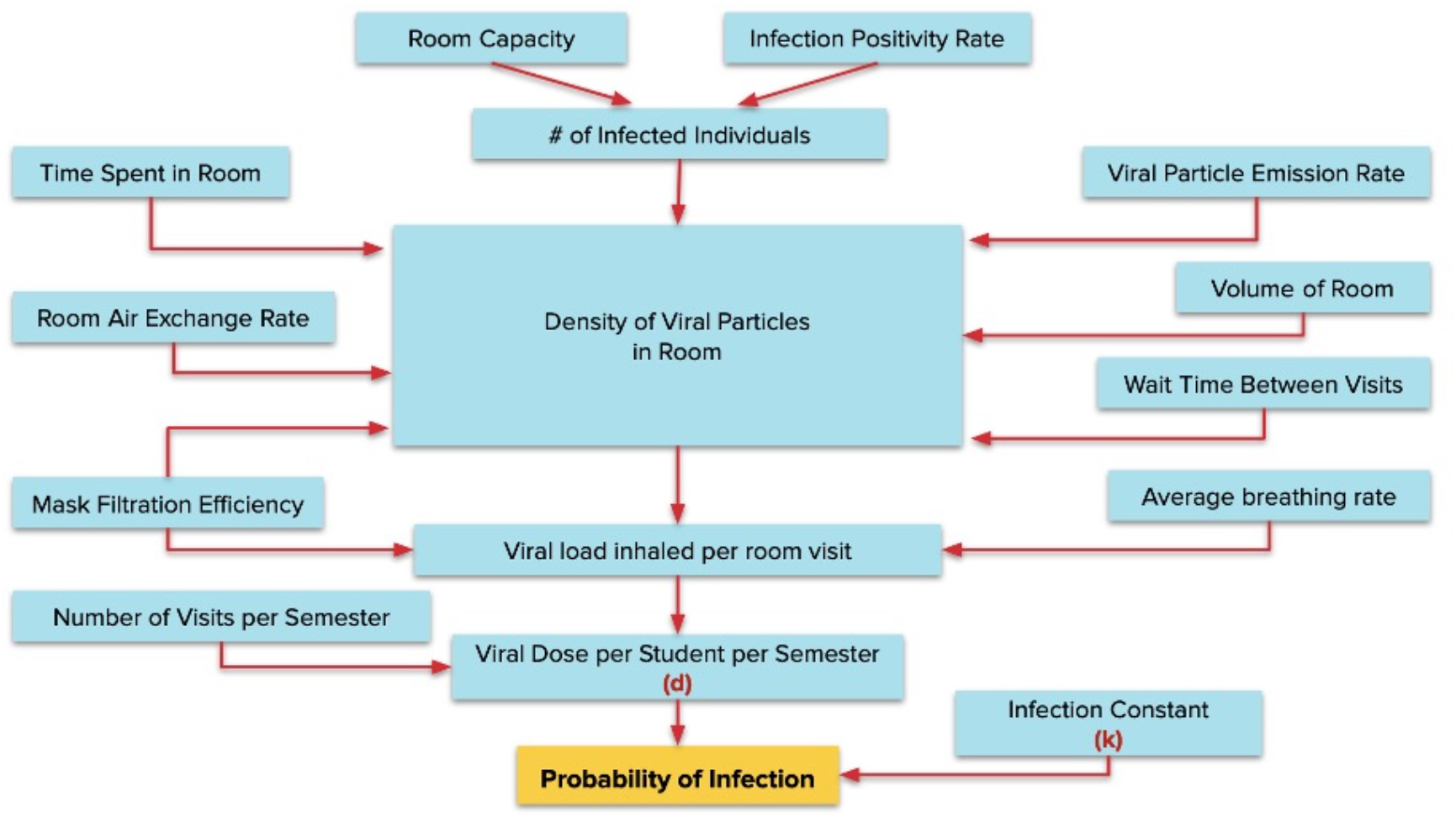
General flowchart of steps in calculating probabilities of infection in university spaces.

#### Teaching Spaces

In-person classes in both large lecture halls and smaller classrooms were analyzed. Input parameters include:

1. University’s COVID-19 positivity rate, *R*_*P*_
2. Normal seating capacity, *N*_*S*_
3. Face mask filtration efficiency, *F*
4. Air exchange rate (hr^-1^), *r*
5. Teaching space volume (L), *V*
6. Class length (min), *t*_*f*_
7. Number of classes attended in that space over the semester, *N*_*Cls*_

Assuming students follow 6 feet social distancing guidelines and each seat is approximately 2 feet x 2 feet, the maximum number of students in the space (*N*_*Stu*_) is:

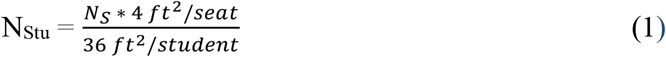

Assuming the number of infected students is proportional to the university’s positivity rate, the number of infected students in the space (*N*_*Inf*_) is:

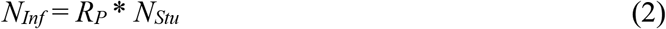

Assuming students wear the designated face mask at all times during the class, the amount of dilution of particles in the air, i.e. the dilution factor, due to the mask (*D*_*mask*_) is:

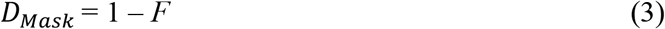

The dilution factor due to HVAC system (*D*_*HVAC*_) is:

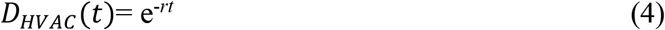

where *t* is the time point of measurement (hr) and *r* is the air exchange rate of the space. (Augenbraun et al., 2020).

Assuming perfect mixing of particles in the total air in the room volume, the increase in viral particle density (*Δ*ρ) in air each minute is:

\# of viral particles exhaled each minute = *N*_*VP*_*= N*_*Inf*_ * 70 particles

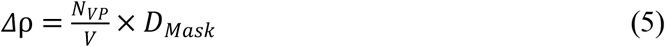

At each time point *t*, up to the duration of one class period, the local density *n(t)* of viral particles (particles/L) in the air inhaled by a healthy individual in the teaching space is incremented by a factor proportional to *Δ*ρ, *D*_*;mask*,_and *D*_*HVAC*_.

For example, if measuring the local density *n(t)* in the air inhaled at the time point of 3 minutes, assuming each minute is a discrete time point, viral density from *t* = 1 minute has been diluted by the HVAC system for 3 minutes, viral density from *t* = 2 minutes has been diluted for 2 minutes, and viral density from *t* = 3 minutes has been diluted for 1 minute. Therefore:

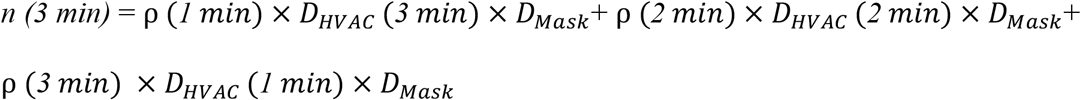

Since each time is being treated as a discrete interval, ρ*(1 min)*, ρ*(2 min)* and ρ*(3 min)* all equal *Δ*ρ, resulting in the relationship:

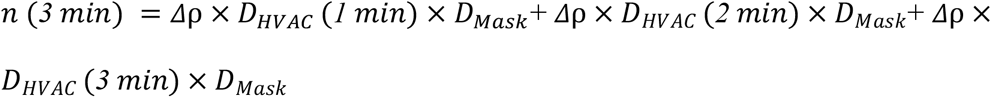

Thus, the local density *n(t)* in inhaled air at time point *t* is determined. Using the volume of air inhaled per minute, 7.5 L/min (derived from volume inhaled per hour, 450 L/min), the viral dose inhaled each minute is determined. The summation of the viral doses inhaled during each minute gives the total viral dose inhaled over the class period:

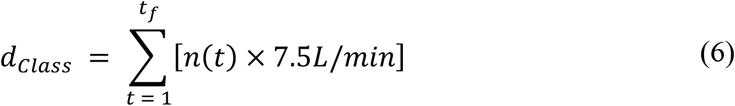

Then, the viral dose inhaled over the semester is:

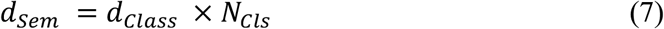

Finally, probability of infection, as described by Watanabe et al. (2010) is:

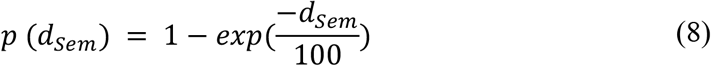

To calculate required wait times between classes held in the same teaching space, the following process was utilized:

The contribution to infection probability made by the viral density from the previous class must be negligible to avoid additive effects for students in the next class. Therefore, the threshold for the maximum probability of infection from one class to the next over the semester was set as 0.1%.

The following parameters are first defined:

Final viral density in the air at the end of this class: ρ_*F*_

Viral density in the air at the start of the next class: ρ_*NC*_

The local density of residual viral particles from the last class (particles/L) in air inhaled by a healthy individual during the next class, *n*_*NC*_ *(t)*, is:

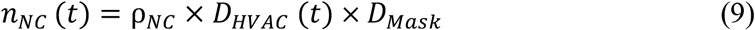

The residual viral dose inhaled by a healthy individual in the next class, *d*_*NC*_, is:

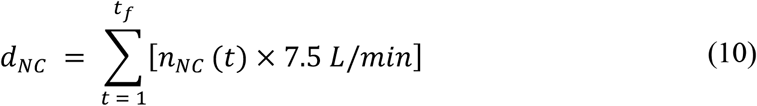

Using this information and the previously described procedure for calculating infection probability, a value for ρ_*NC*_ that produces a probability of infection from the next class over the semester that is < 0.1% can be found.

Finally, the required wait time (*τ*) for the viral density to dilute from ρ_*F*_ to ρ_*NC*_ is:

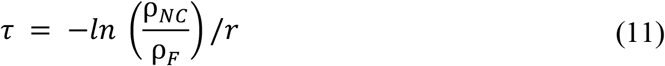

where *r* is the air exchange rate of the space.

#### Dining Halls

For dining halls, both in-person dining and meal pick-up scenarios were analyzed. The approach taken was very similar to that of teaching spaces. Input parameters include:

1. University’s COVID-19 positivity rate, *R*_*P*_
2. Normal seating capacity, *N*_*S*_
3. Filtration efficiency of face masks worn (if any), *F*
4. Air exchange rate (hr^-1^), *r*
5. Dining hall volume (L), *V*
6. Amount of time spent in dining hall per meal (min), *t*_*f*_
7. Number of weeks in the semester, *N*_*Wks*_

Furthermore, it was assumed that students eat 3 meals per day, 7 days per week, and that face masks are not worn in the in-person dining scenario. In the meal pick-up scenario, it was assumed that students remain 6 feet distanced at all times, adhere to the length of meal pick-up windows, and do not pause to interact with others during the meal pick-up.

With these parameters and assumptions, the same approach used for teaching spaces was implemented to calculate infection probabilities for in-person dining and meal pick-ups, as well as required wait times between meal pick-up batches.

#### Residential spaces

Dorm rooms and bathrooms, two of the largest sources of interaction between students in residential buildings, were analyzed.

#### Dorm Rooms

Input parameters include:

1. Number of students sharing the dorm room, *N*_*Stu*_
2. Air exchange rate (hr^-1^), *r*
3. Dorm room volume (L), *V*
4. Amount of time spent in the dorm room per day, *t*_*f*_

It was assumed that face masks are not worn within the dorm room. The infection probability was calculated over a 2-week period rather than the entire semester as was done for the other spaces. This was done to cover the typical 2-week infection period of an individual with COVID-19.

With these parameters and assumptions, the same approach to calculating infection probability that was used for teaching spaces was implemented.

#### Bathrooms

Input parameters include:

1. Number of students per dorm room, *N*_*Stu*_
2. Number of dorm rooms sharing the bathroom, *N*_*Rms*_
3. Air exchange rate (hr^-1^), *r*
4. Bathroom volume (L), *V*
5. Amount of time per bathroom use (min), *t*_*B*_
6. Amount of time per shower use (min), *t*_*S*_
7. Wait time between bathroom uses (min), *τ*_*WB*_
8. Wait time between shower uses (min), *τ*_*WS*_
9. Number of bathroom uses per day, *N*_*B*_
10. Number of shower uses per day, *N*_*S*_

It is important to note that unlike other university spaces, bathroom wait times are an input parameter rather than a calculated output parameter. This is because wait times between bathroom uses and shower uses are interrelated, i.e. if the wait time between shower uses is lengthened, the wait time between bathroom uses may be shortened safely. Therefore, the exact balance between these parameters must be a user decision rather than a calculated output.

It was assumed that face masks are worn at all times in the bathroom, except during shower use. It was also assumed that showers are located in the bathroom. The infection probability was again calculated over a 2-week period rather than the entire semester to cover the typical 2-week infection period of an individual with COVID-19.

With these parameters and assumptions, a similar approach to that used for teaching spaces was used to calculate the infection probability.

The final density ρ_*f*_ of viral particles in the air at the end of a bathroom use by an infected student is calculated using the parameters, *Δ*ρ, *D*_*HVAC*_ *(t)*, and *D*_*mask*_.

The diluted density ρ_*Dil*_ after the wait time between bathroom uses is:

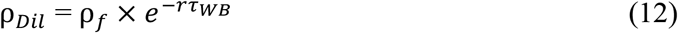

During the next bathroom use by an uninfected student, the local density *n(t)* of viral particles (particles/L) at time *t* in air inhaled by the uninfected individual is:

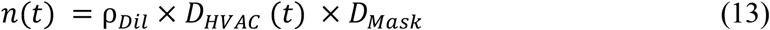

The viral dose, *d*_*B*_, during this bathroom visit is again calculated as:

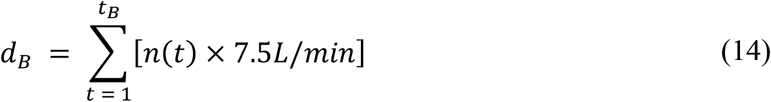

This process can be repeated for shower uses with duration (*t*_*S*_) and wait time (*τ*_*WS*_). In this case, *D*_*Mask*_would not be included, as mask-wearing while showering is unfeasible. The viral dose during a shower visit would be *d*_*S*_.

To calculate the dose inhaled over the infection period:

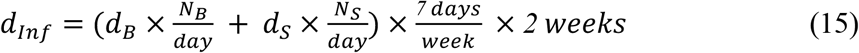

Finally, to calculate infection probability:

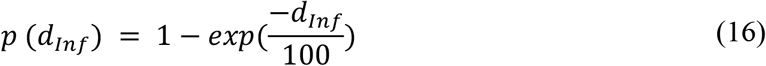

### Probabilities of Infection in Specific University Spaces

These approaches were applied to specific teaching spaces, dining halls, and residential spaces in the Harvard College Cambridge campus, Harvard University Allston campus, and Stanford University campus. Data regarding air exchange rates, seating capacities, and room dimensions was collected for various spaces in these university campuses.

There is more variability in air flow in small classrooms due to doors/windows opening, foot traffic, etc. For this reason, it was conservatively estimated that these spaces have air exchange rates similar to those of office spaces, i.e. 3 air exchanges per hour (Augenbraun et al., 2020). For large lecture halls and dining halls, however, air exchange rates were collected and used for calculations.

Two extremes of university-wide COVID-19 positivity rates were tested. A low infection rate (LIR) case was tested, with a 0.1% positivity rate, as well as a high infection rate (HIR) case, with a 1% positivity rate.

Since exact numerical results depend heavily on scenario-dependent variables, a few example scenarios in the university spaces tested (i.e. teaching spaces, dining halls, and residential spaces) were selected. The specific scenarios to which this model was applied in different universities are:

Teaching spaces:

1. 1.5 hours per class
2. 180 classes in the space over the semester

Dining Halls:

#### In-person dining

1. 1 hour per meal
2. 3 meals per day
3. Eating in dining hall 7 days per week
4. 12 weeks per semester

#### Meal pick-ups

1. 10 minutes per meal pick-up
2. 3 meal pick-ups per day
3. Picking up meals 7 days per week
4. 12 weeks per semester

Residential spaces

#### Dorm rooms

1. 2 students per room
2. 1 student is infected
3. Spending 18 hours per day in the dorm room
4. Living in dorm room 7 days per week

#### Bathrooms

1. 3 dorm rooms with 2 students each share a bathroom
2. 1 dorm room is infected
3. 5 minutes per bathroom use
4. 20 minutes per shower use
5. 40-minute wait time between bathroom uses by students from different rooms
6. 80-minute wait time between shower uses by students from different rooms
7. 4 bathroom uses per day
8. 1 shower use per day
9. Using these residential bathrooms 7 days per week

### Evaluation of Impact of Face Masks

The risk assessment model estimates the infection probability. By setting the maximum risk of infection as 1%, this model can be used to evaluate the ability of a face mask to sufficiently mitigate infection risk. Filtration efficiencies for these masks were assigned based on the usual range of values observed (Hao et al., 2020; Konda et al., 2020).

1. Procedural mask with filtration efficiency of 70%
2. Cotton mask with filtration efficiency of 45%
3. Cotton mask with filtration efficiency of 15%
4. N95 respirator with filtration efficiency of 95%
5. KN95 mask with filtration efficiency of 90%

### Web Application Development

A web application implementing the mathematical model was deployed using Amazon Web Services. The web application was developed using HTML, Javascript, and Python. This website was designed to be user-friendly and efficient in obtaining infection probabilities for different university spaces (Figure 2). It also provides users with CDC information and university policies aimed at mitigating viral transmission on campus (Figure 2). Like the algorithm described, conservative assumptions were used to output infection probabilities. The COVID-19 dashboard for each university, in which student, faculty, and staff infections are tracked, was either embedded or linked in the web application to allow users to input their universities’ most up-to-date positivity rates.

**Figure 2:**
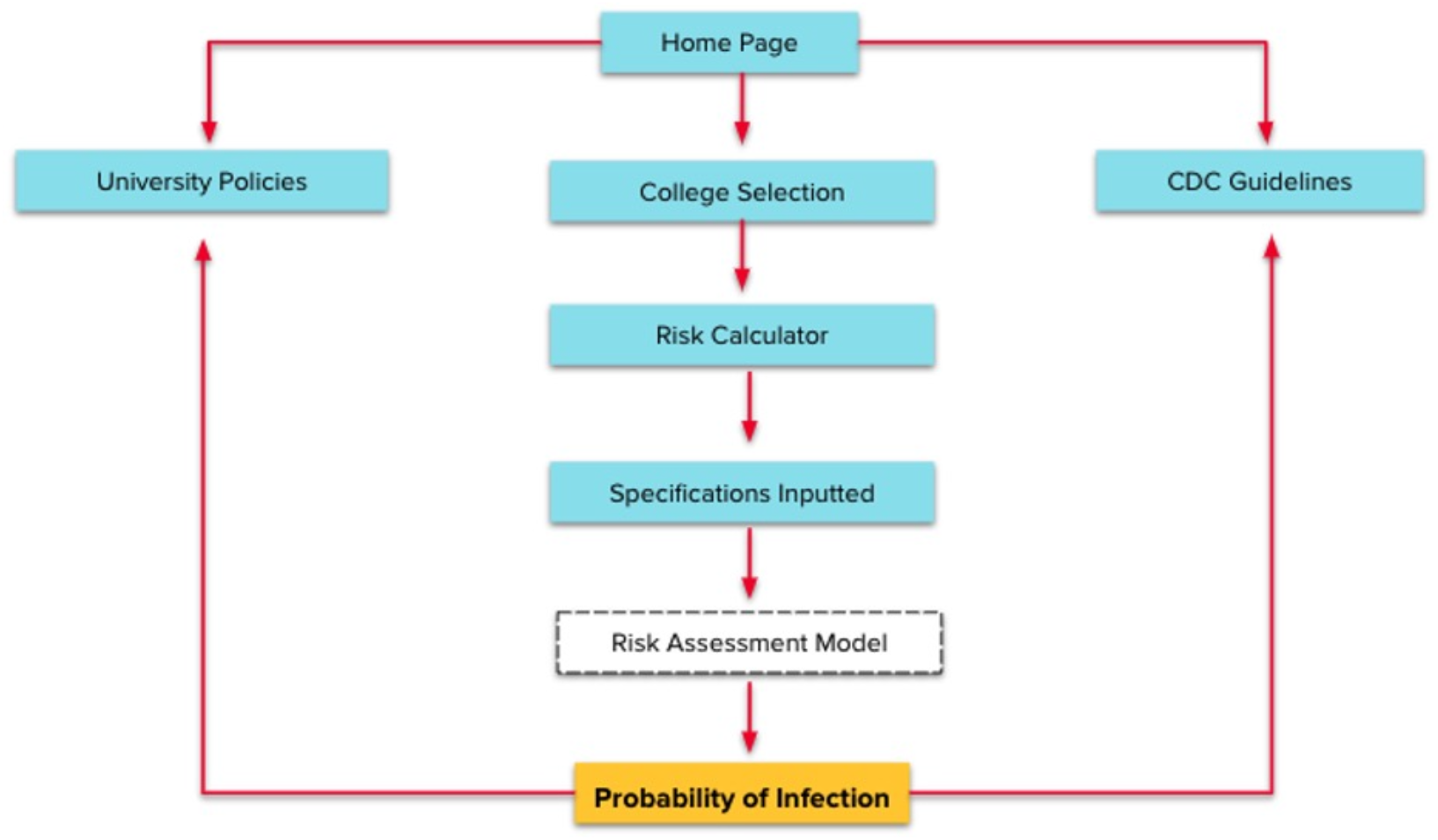
General flow chart of web-application layout.

To ensure the validity of the web application’s algorithm, cross-data comparison between the results of the web application algorithm and manual calculations through the approaches detailed previously were continuously performed throughout the development of the web application.

## RESULTS

### Model Development

The model to calculate infection probability is based on a previously developed, exponential dose-response model of infection probability for SARS coronavirus, which has recently been used to calculate infection probability with SARS-CoV-2 (Watanabe et al., 2010; Augenbraun et al., 2020). This model defines infection probability as:

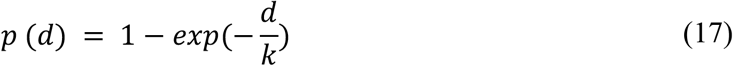

where *d* is the viral dose and *k* is the infection constant for the respective virus (Watanabe et al., 2010; Augenbraun et al., 2020)

To calculate viral dose, the local density of viral copies in air inhaled by an individual must be determined (Augenbraun et al., 2020). On average, an individual infected with SARS-CoV-2 exhales 35-70 viral particles/minute (Augenbraun et al., 2020). By the time the viral particles spread and reach a healthy individual, there is a specific local density *n* of viral particles (particles/L) in the inhaled air. The inhaled viral dose is then calculated as:

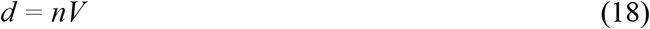

where the inhaled viral dose (*d*) is a product of the local viral density (*n*) in inhaled air and the volume of air inhaled (V).

Factors that dilute viral density are face mask usage, HVAC filtration systems, and elapsed time, making it essential to quantify their effect on the local density *n* of viral particles in inhaled air. In the case of face masks, the dilution factor *D*_*Mask*_ is the degree of viral density dilution that results from passage through the mask. Therefore, *D*_*Mask*_ is simply the fraction of viral particles that pass through the mask and is defined as:

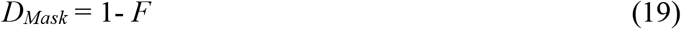

where the dilution factor *(D*_*Mask*_) is a function of the mask filtration efficiency (*F*) for particles *≥* 0.3 microns in size.

For HVAC filtration systems, when low flow rates result in a mixing of contaminated and fresh air, the dilution factor *D*_*HVAC*_ follows the relationship:

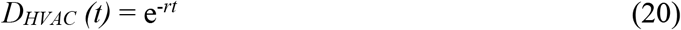

where the airborne viral particle dilution factor (*D*_*HVAC*_) is dependent on the number of air exchanges per hour (*r*) and the duration of time in hours (*t*). (Augenbraun et al., 2020).

The general approach to calculating infection probability in a desired setting was based on the assumption that the number of infected individuals sharing a particular space is representative of the university’s infection positivity rate. The number of viral particles emitted was calculated using the assumed number of infected individuals and a viral particle exhalation rate of 70 particles/minute. The filtration efficiency of the face mask worn, if any, was used to calculate *D*_*Mask*_, and the room’s air exchange rate was used to calculate *D*_*HVAC*_, which in turn were used, along with the room volume, to calculate the local density of viral particles, *n*, in inhaled air. Using the average breathing rate of 450 L/hr, the viral dose inhaled in a given amount of time was calculated. This viral dose and an infection constant of *k* = 100 (i.e. each inhaled viral particle contributes 1% infection probability) were used to calculate infection probability.

### Analysis Using Risk Assessment Model for Generalized University Spaces

The model was first applied to generalized university spaces. Values for different space parameters (positivity rate, room volume, air exchange rate, mask filtration efficiency, normal room capacity, time spent in the space per visit, and total number of visits to the space over a semester) were selected such that they fall within the range of reasonable values for university spaces while still providing wide variation. Thus, five generic university spaces were created (S1 Table), to which the model was applied. In general, mask filtration efficiency, air exchange rate, room capacity, and campus COVID-19 positivity rate were dominant parameters influencing infection probability (Figure 3).

**Figure 3:**
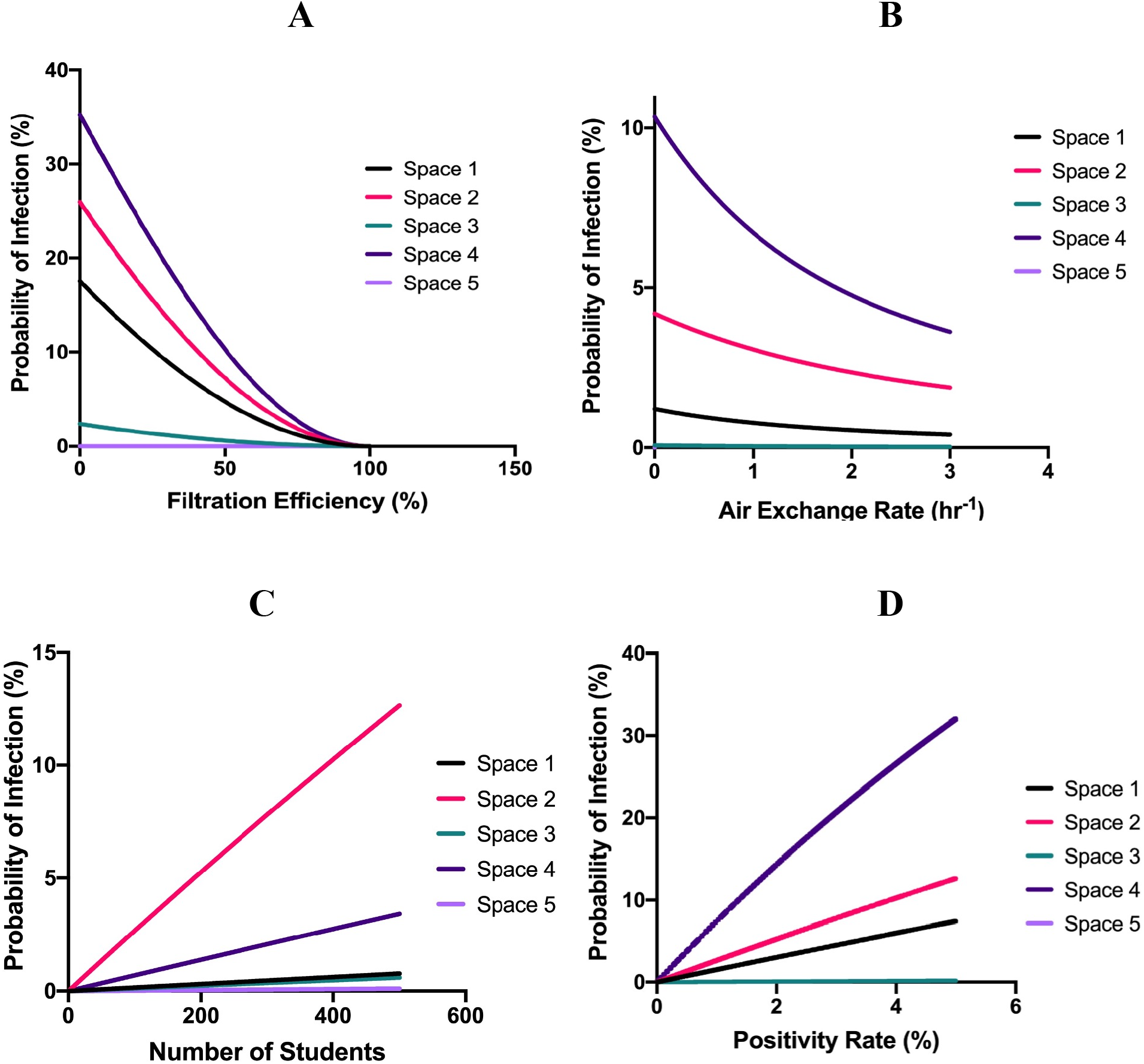
Relationships between dominant parameters in the model and probability of infection in five generic university spaces. **A** - Mask Filtration Efficiency vs. Probability of Infection, assuming constant values for all other parameters (i.e., positivity rate, room volume, air exchange rate, normal room capacity, time per visit, and total number of visits per semester) in each generic space. **B** - Air Exchange Rate vs. Probability of Infection, assuming the same constant values for all other parameters in each generic space. **C** - Number of Students vs. Probability of Infection, assuming the same constant values for all other parameters in each generic space. **D** - Positivity Rate vs. Probability of Infection, assuming the same constant values for all other parameters in each generic space. (See S1 Table for details on these parameters for these five generic spaces).

In spaces that are used sequentially by different groups of students (e.g., consecutive classes in teaching spaces, meal pick-ups in dining halls, bathroom uses) the infection probability during the next use of the space over a semester was determined, through application of the model, to depend on many of the same parameters. However, wait time between uses also plays an important role (Figure 4).

**Figure 4:**
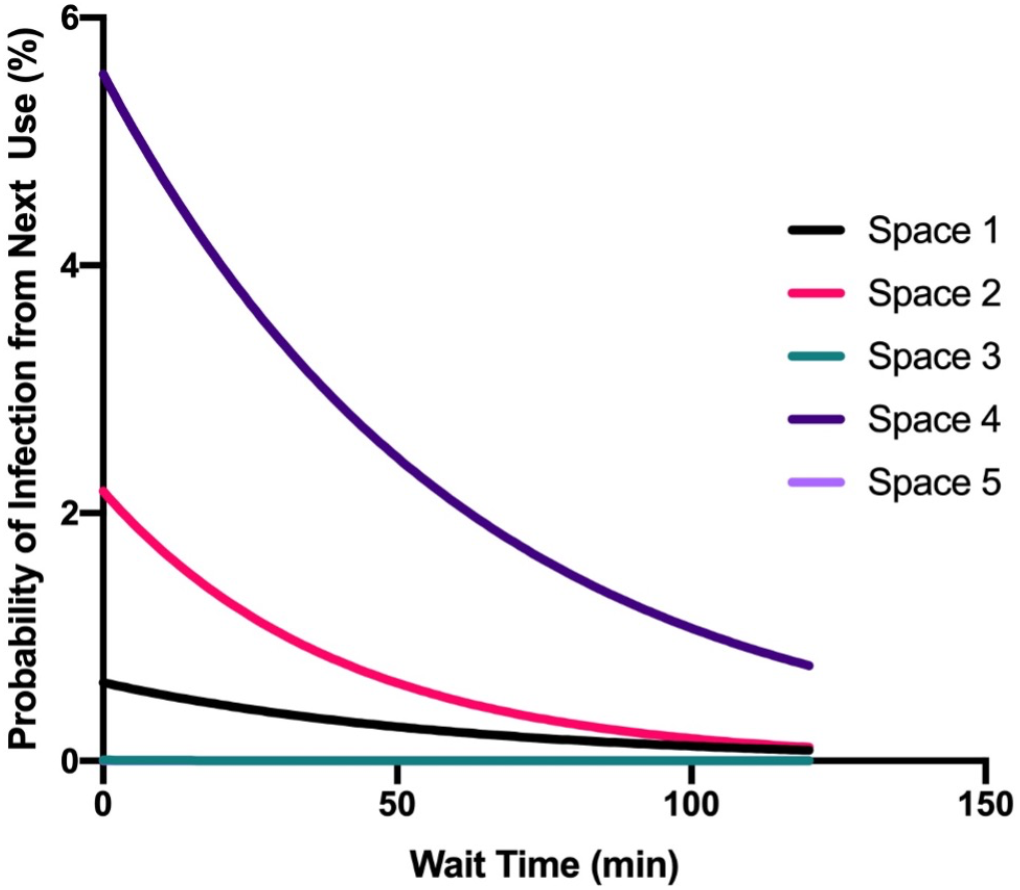
Relationship between wait time in between uses and probability of infection due to the subsequent use of the same space in five generic university spaces (See S1 Table for details of spaces). These calculations assumed constant values for all other parameters (i.e., room volume, air exchange rate, face mask filtration efficiency, normal room capacity, time per visit, and total number of visits per semester).

### Analysis Using Risk Assessment Model for Specific University Spaces

The model was applied to specific university spaces and scenarios, using values specific to particular spaces at Harvard and Stanford Universities. Both Low Infection Rate (0.1%) (LIR) and High Infection Rate (1%) (HIR) scenarios were used for all calculations.

#### Required Filtration Efficiency Results

For each university campus, specific parameter value ranges are required to ensure safety (*p* < 1%) and feasibility of reopening spaces (e.g., cannot have exorbitant wait times > 5 minutes between meal pick-up batches to ensure all students receive meals in a timely fashion). To determine the required mask filtration efficiencies, multiple specific spaces of each type (e.g. teaching spaces) in each university were analyzed to determine the minimum filtration efficiency required to ensure safety and feasibility across all spaces of a single type for a given university. For example, the required filtration efficiency for lecture halls at Harvard was determined by testing multiple Harvard lecture halls and finding the single filtration efficiency required to meet the p < 1% criteria for all. Through this analysis, mask filtration efficiencies required to ensure safety and feasibility for each space type in each university were determined (Table 1). Understandably, required filtration efficiencies were greater in the HIR case than the LIR case (Table 1), as there is greater exposure to infected individuals in the HIR case. Differences between required filtration efficiencies across universities and space types occurred due to variations in the space parameters of different universities and space types. For example, required filtration efficiencies for meal pick-ups from dining halls were much lower for the Stanford University campus compared to the Harvard University Cambridge campus (Table 1) due to factors such as greater room volumes in Stanford dining halls. Similarly, required filtration efficiencies were generally lower in small classrooms than large lecture halls (Table 1) due to factors such as lower seating capacities and greater air exchange rates.

**Table 1:**
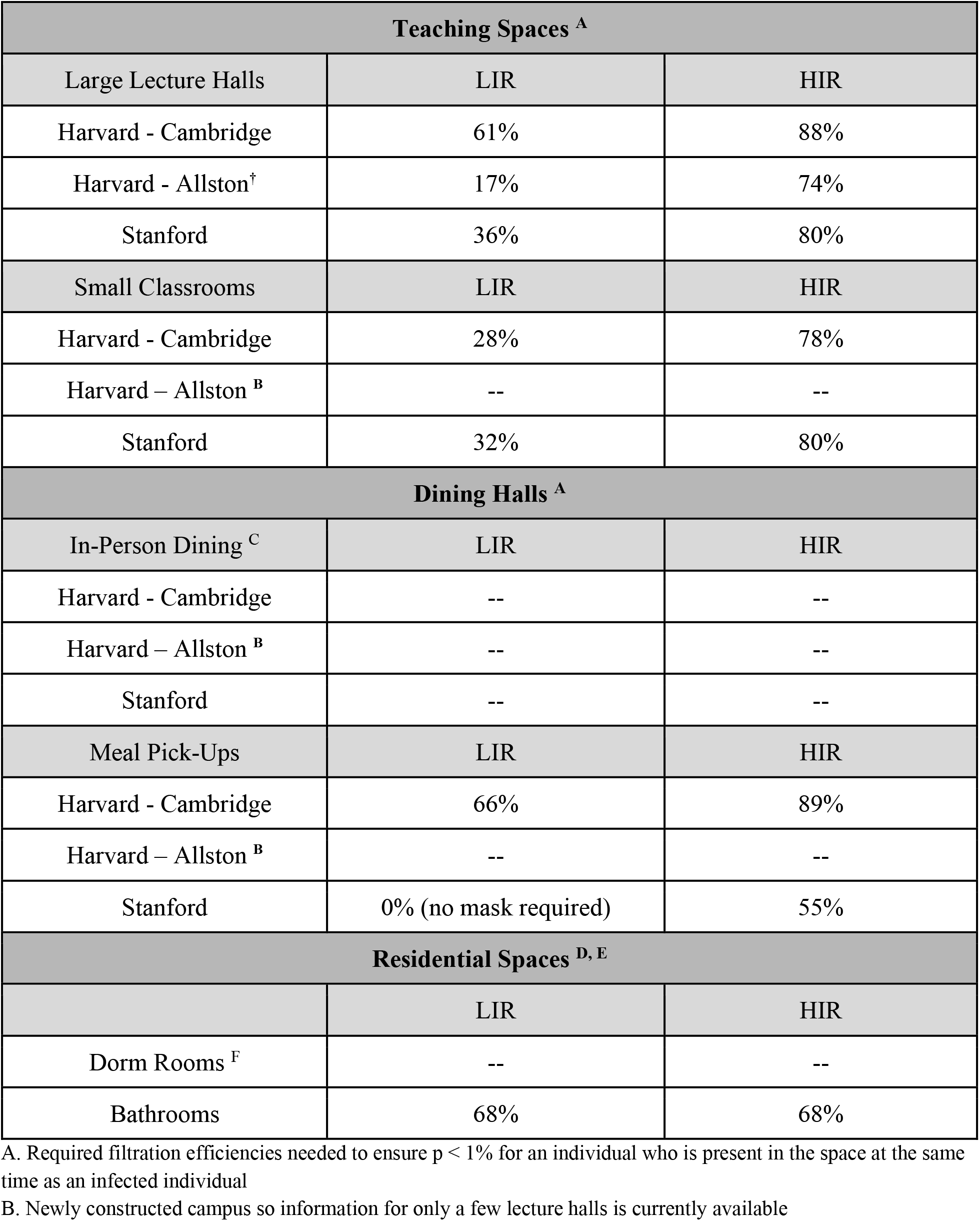
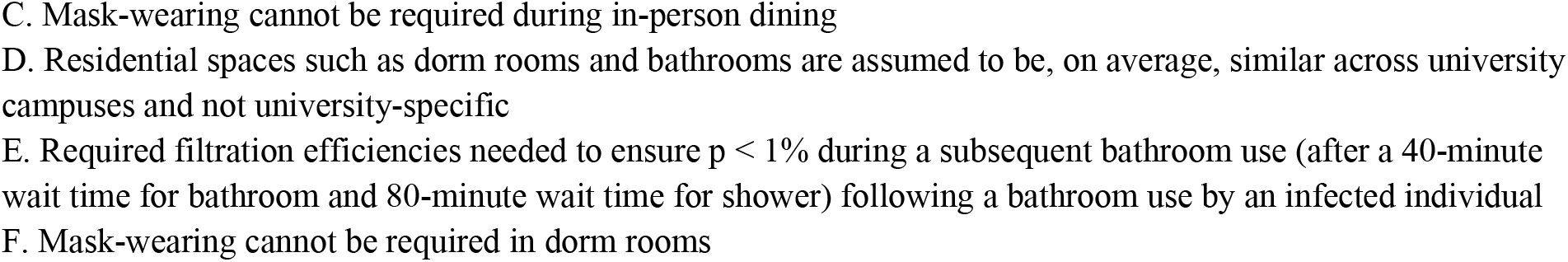
Required face mask filtration efficiencies to yield low probability of infection.

| Teaching Spaces <sup>A</sup> |  |  |
| --- | --- | --- |
| Large Lecture Halls | LIR | HIR |
| Harvard - Cambridge | 61% | 88% |
| Harvard - Allston<sup>†</sup> | 17% | 74% |
| Stanford | 36% | 80% |
| Small Classrooms | LIR | HIR |
| Harvard - Cambridge | 28% | 78% |
| Harvard – Allston <sup>B</sup> | -- | -- |
| Stanford | 32% | 80% |
| Dining Halls <sup>A</sup> |  |  |
| In-Person Dining <sup>C</sup> | LIR | HIR |
| Harvard - Cambridge | -- | -- |
| Harvard – Allston <sup>B</sup> | -- | -- |
| Stanford | -- | -- |
| Meal Pick-Ups | LIR | HIR |
| Harvard - Cambridge | 66% | 89% |
| Harvard – Allston <sup>B</sup> | -- | -- |
| Stanford | 0% (no mask required) | 55% |
| Residential Spaces <sup>D, E</sup> |  |  |
|  | LIR | HIR |
| Dorm Rooms <sup>F</sup> | -- | -- |
| Bathrooms | 68% | 68% |

A. Required filtration efficiencies needed to ensure $p < 1\%$ for an individual who is present in the space at the same time as an infected individual
B. Newly constructed campus so information for only a few lecture halls is currently available
- C. Mask-wearing cannot be required during in-person dining - D. Residential spaces such as dorm rooms and bathrooms are assumed to be, on average, similar across university campuses and not university-specific - E. Required filtration efficiencies needed to ensure $p < 1\%$ during a subsequent bathroom use (after a 40-minute wait time for bathroom and 80-minute wait time for shower) following a bathroom use by an infected individual - F. Mask-wearing cannot be required in dorm rooms

It is important to note that these were the required filtration efficiencies to ensure that the infection risk at the time of maximum exposure is <1% over the relevant time period. For example, for teaching spaces and meal pick-ups, Table 1 presents the necessary filtration efficiencies for ensuring that an individual present in the space at the same time as an infected individual will have <1% infection probability over a semester. For bathrooms, since individuals from different rooms were assumed to not be permitted to be present at the same time, maximum exposure occurs during the subsequent visit (after the constant wait times previously defined) following a bathroom visit by individuals from an infected room. Therefore, filtration efficiencies in Table 1 for bathrooms are those that are necessary during subsequent visits.

#### Efficacy of Existing Face Masks

Comparison of the performance of existing masks relative to these required filtration efficiencies allowed for the determination of their ability to successfully maintain an infection probability <1% over the relevant time period in these specific university spaces and aforementioned scenarios. The N95 respirator and KN95 mask have high filtration efficiencies and therefore were found to be acceptable across all the spaces and university campuses (Table 2). Typical procedural masks have sufficient filtration properties to maintain p < 1% across all the spaces when infection rates are low but not when they are high (Table 2). The cotton mask with 15% filtration efficiency was found to provide insufficient protection in almost all spaces and university campuses (Table 2).

**Table 2:** Evaluation of efficacy of existing face masks in mitigating risk.

|  | Procedural Mask (70%) |  | Cotton Mask (45%) |  | Cotton Mask (15%) |  | N95 Respirator (95%) |  | KN95 Mask (90%) |  |
| --- | --- | --- | --- | --- | --- | --- | --- | --- | --- | --- |
| Teaching spaces |  |  |  |  |  |  |  |  |  |  |
| Large Lecture Halls | LIR | HIR | LIR | HIR | LIR | HIR | LIR | HIR | LIR | HIR |
| Harvard - Cambridge | ✓ | ✗ | ✗ | ✗ | ✗ | ✗ | ✓ | ✓ | ✓ | ✓ |
| Harvard - Allston | ✓ | ✗ | ✓ | ✗ | ✗ | ✗ | ✓ | ✓ | ✓ | ✓ |
| Stanford | ✓ | ✗ | ✓ | ✗ | ✗ | ✗ | ✓ | ✓ | ✓ | ✓ |
| Small Classrooms | LIR | HIR | LIR | HIR | LIR | HIR | LIR | HIR | LIR | HIR |
| Harvard - Cambridge | ✓ | ✗ | ✓ | ✗ | ✗ | ✗ | ✓ | ✓ | ✓ | ✓ |
| Harvard - Allston | -- | -- | -- | -- | -- | -- | -- | -- | -- | -- |
| Stanford | ✓ | ✗ | ✓ | ✗ | ✗ | ✗ | ✓ | ✓ | ✓ | ✓ |
| Dining Halls |  |  |  |  |  |  |  |  |  |  |
| Meal Pick-Ups | LIR | HIR | LIR | HIR | LIR | HIR | LIR | HIR | LIR | HIR |
| Harvard - Cambridge | ✓ | ✗ | ✗ | ✗ | ✗ | ✗ | ✓ | ✓ | ✓ | ✓ |
| Harvard - Allston | -- | -- | -- | -- | -- | -- | -- | -- | -- | -- |
| Stanford | ✓ | ✓ | ✓ | ✗ | ✓ | ✗ | ✓ | ✓ | ✓ | ✓ |
| Residential Spaces |  |  |  |  |  |  |  |  |  |  |
|  | LIR | HIR | LIR | HIR | LIR | HIR | LIR | HIR | LIR | HIR |
| Bathrooms | ✓ | ✓ | ✗ | ✗ | ✗ | ✗ | ✓ | ✓ | ✓ | ✓ |

#### Other Output Parameters

Assuming the calculated values for required mask filtration efficiencies are enforced, specific wait times and infection probabilities were calculated for each university space (Tables 3-5). Infection probabilities were, as expected, <1% in all spaces where masks with the required filtration efficiency listed in Table 1 for the given space type are worn (Tables 3-5). However, due to variations between specific spaces, the magnitude of the infection probability differs between spaces (Tables 3-5). Therefore, while Table 1 provides the filtration efficiencies required to ensure p < 1% over the relevant time period across spaces of a given type in each university, Tables 3-5 provide the specific output parameter values for each individual space.

**Table 3:** Output parameter results for teaching spaces when using the required filtration efficiencies defined in Table 1.

|  | Original Room Capacity | Room Capacity Following 6 ft Distancing | Wait time between classes <sup>A</sup> |  | Probability of infection <sup>B</sup> |  |
| --- | --- | --- | --- | --- | --- | --- |
| Harvard - Cambridge |  |  |  |  |  |  |
| Lecture Halls |  |  | LIR | HIR | LIR | HIR |
| Science Center Hall B | 492 | 54 | 1.27 hours | 1.46 hours | 0.50% | 0.47% |
| Science Center Hall E | 131 | 14 | 2.84 hours | 2.96 hours | 0.99% | 0.93% |
| Sanders Theater | 1000 | 111 | 0.46 hours | 0.63 hours | 0.22% | 0.20% |
| Small Classrooms |  |  | LIR | HIR | LIR | HIR |
| Science Center Room 105 | 26 | 2 | 0.33 hours | 0.47 hours | 0.91% | 0.86% |
| Maxwell Dworkin Seminar Room | 10 | 1 | 0.35 hours | 0.49 hours | 0.99% | 0.91% |
| Sever Hall Seminar Room | 21 | 2 | 0.27 hours | 0.41% | 0.77% | 0.72% |
| Harvard - Allston |  |  |  |  |  |  |
| Lecture Halls |  |  | LIR | HIR | LIR | HIR |
| SEC 1.402 | 60 | 6 | 0.49 hours | 0.47 hours | 0.81% | 0.79% |
| SEC LL.224 | 100 | 11 | 0.82 hours | 0.81 hours | 0.99% | 0.97% |
| Stanford |  |  |  |  |  |  |
| Lecture Halls |  |  | LIR | HIR | LIR | HIR |
| Large Hall | 135 | 15 | 1.72 hours | 1.65 hours | 0.99% | 0.97% |
| Medium Hall | 54 | 6 | 0.49 hours | 0.66 hours | 0.99% | 0.97% |
| Small Classrooms |  |  | LIR | HIR | LIR | HIR |
| Seminar Room | 16 | 1 | 1.01 hours | 0.94 hours | 0.99% | 0.84% |

A. Wait times that are required to ensure safety in the class subsequent to one in which an infected individual was present
B. Probabilities of infection over a semester for individuals who are attending a class with an infected individual, i.e. they are present in the space at the same time as the infected individual

In the case of teaching spaces, new room capacities were derived from original capacities to follow 6 feet social distancing in the three university campuses. Using these capacities and the filtration efficiencies for each case listed in Table 1, infection probabilities over a semester in these teaching spaces (Table 3) were calculated. It is important to note that these probabilities were obtained for the case in which infected individuals are present in the teaching space at the same time as healthy individuals. These probabilities are independent of wait times between classes (Table 3), which are necessary to maintain acceptably low infection probabilities for individuals attending subsequent classes in the teaching spaces.

For dining halls, room capacities following 6 feet social distancing were again determined from original capacities for specific dining halls in Harvard and Stanford universities. Using these capacities and the filtration efficiencies for each case listed in Table 1, infection probabilities over a semester in these dining halls (Table 4) were calculated both for in-person dining and meal pick-up scenarios previously described. Notably, infection probabilities for in-person dining were frequently >1% (Table 4) because mask usage cannot be enforced during in-person dining. In contrast, infection probabilities for meal pick-ups were significantly <1% (Table 4), largely due to mask use and the short duration spent in the dining hall during the pick-up. It is also important to note that these probabilities are once again for the case in which infected individuals are present in the dining hall at the same time as healthy individuals. These probabilities are independent of wait times between meal batches (Table 4), which are necessary to maintain acceptably low infection probabilities for individuals picking up meals in subsequent batches.

**Table 4:** Output parameter results for dining halls when using the required filtration efficiencies defined in Table 1.

|  | Original Room Capacity | Room Capacity Following 6 ft Distancing | Wait time in between batches <sup>A, B</sup> |  | Probability of infection <sup>C</sup> |  |
| --- | --- | --- | --- | --- | --- | --- |
| Harvard - Cambridge |  |  |  |  |  |  |
| In-person dining |  |  | LIR | HIR | LIR | HIR |
| Annenberg Dining Hall | 800 | 88 | -- | -- | 2.00% | 18.34% |
| Dunster Dining Hall | 250 | 27 |  |  | 7.14% | 52.28% |
| Kirkland Dining Hall | 200 | 22 |  |  | 13.75% | 77.22% |
| Meal pick-ups |  |  | LIR | HIR | LIR | HIR |
| Annenberg Dining Hall | 800 | 88 | 10-30 seconds | 1-3 minutes | 0.01% | 0.01% |
| Dunster Dining Hall | 250 | 27 |  |  | 0.03% | 0.03% |
| Kirkland Dining Hall | 200 | 22 |  |  | 0.05% | 0.06% |
| Stanford |  |  |  |  |  |  |
| In-person dining |  |  | LIR | HIR | LIR | HIR |
| Arrillaga Family Dining Commons | 675 | 75 | -- | -- | 0.73% | 6.95% |
| Manzanita Dining Hall | 320 | 35 |  |  | 0.69% | 6.60% |
| Ricker Dining Hall | 292 | 32 |  |  | 0.63% | 6.13% |
| Meal pick-ups |  |  | LIR | HIR | LIR | HIR |
| Arrillaga Family Dining Commons | 675 | 75 | 10-30 seconds | 10-30 seconds | 0.02% | 0.05% |
| Manzanita Dining Hall | 320 | 35 |  |  | 0.02% | 0.05% |
| Ricker Dining Hall | 292 | 32 |  |  | 0.02% | 0.04% |

A. Magnitude of wait times is much smaller for meal pick-ups so they are provided in order of magnitude ranges, which likely carry greater utility for guiding university policies
B. Wait times required to ensure safety in a meal pick-up batch subsequent to one in which an infected individual was present
C. Probabilities of infection over a semester for individuals in the same meal pick-up batch as an infected individual

For residential spaces, using the required filtration efficiency listed in Table 1 and variables specified by the scenarios previously described, infection probabilities over a 2-week infection period were calculated for typical dorm rooms and bathrooms of two different sizes and levels of ventilation (Table 5). For bathrooms, individuals from different rooms were assumed to never be present at the same time. Unlike teaching spaces and dining halls, infection probabilities in bathrooms were dependent on the wait time between uses. For this reason, infection probabilities for bathrooms listed in Table 5 were calculated for the constant wait times previously defined (40 minutes between bathroom uses and 80 minutes between shower uses).

**Table 5:** Output parameter results for residential spaces when using the required filtration efficiencies defined in Table 1.

| Dorm Rooms |  |  |
| --- | --- | --- |
|  |  | Probability of infection |
| Dorm Room |  | 100% |
| Bathrooms |  |  |
| Bathroom Size | Ventilation | Probability of infection <sup>A, B</sup> |
| Large (3000 ft<sup>3</sup>) | High (5 exchanges/hr) | 0.06 % |
|  | Low (3 exchanges/hr) | 0.66 % |
| Small (2000 ft<sup>3</sup>) | High (5 exchanges/hr) | 0.09 % |
|  | Low (3 exchanges/hr) | 0.99 % |

A. For the worst-case scenario in which both students from the infected room are present in the bathroom at the same time
B. Calculated when using the constant wait times previously defined (40 minutes for bathroom uses, 80 minutes for shower uses)

### Web Application Based on Risk Assessment Model

The web application allows users to calculate infection probabilities in teaching spaces in a way that is tailored to their settings. The user is required to input the space dimensions, the length of time inside the space per class, and the number of times they will attend this class over the semester (Figure 5). A user can also input the filtration efficiency of their mask and the air exchange rate (Figure 5). However, if the user does not know or fails to input either, a base-case scenario is calculated. This calculation assumes a mask filtration efficiency of 70%, the filtration efficiency of a surgical mask (Hao et al., 2020), and an air exchange rate of 1.2 cycles per hour. While the model was initially used to identify conditions necessary to maintain <1% infection risk, the web application allows more creativity by allowing conditions to be input by the user and producing an infection probability based on the inputs (Figure 5).

**Figure 5:**
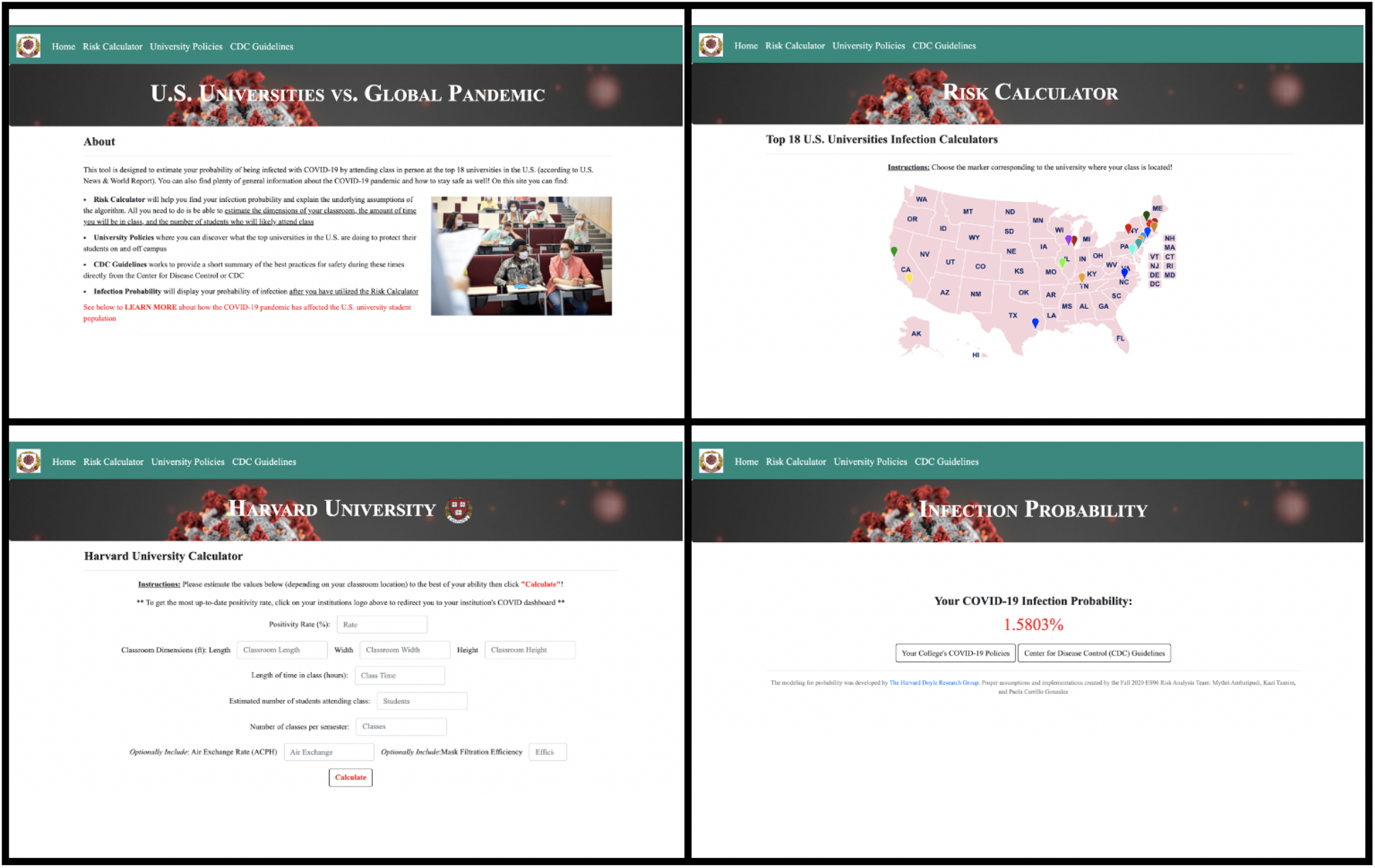
Screenshots of web application predicting the probability of infection of SARS-CoV-2. (https://infection-detection.herokuapp.com/).

## DISCUSSION

A model for quantifying SARS-CoV-2 infection risk in university spaces has been created. Its application to teaching, dining, and residential spaces can provide critical information regarding safety in university facilities and guide university mandates to increase safety. The web application provides the added functionality for any user to determine infection risk in a setting.

This model and tool were formulated using several assumptions. An infection probability <1% over a semester was assumed to be a reasonable threshold for acceptable risk, similar to Augenbraun et al. (2020). At this threshold, the risk of death due to COVID-19 is smaller than the all-cause mortality rate for the university-age demographic (Augenbraun et al., 2020). It was presumed that the number of infected students is proportional to the positivity rate. This yields fractional numbers of infected individuals, which is mathematically equivalent to whole numbers of infected individuals present for a fraction of the duration. The model included the assumption of perfect mixing of particles in a space, which is more appropriate in small spaces. It is likely not feasible to calculate a person-by-person risk assessment based on an individual’s location relative to an infected person. The approach to the HVAC dilution factor (*D*_*HVAC*_) assumes low air flow rates that cause mixing of fresh and contaminated air, which is especially likely in larger spaces with inefficient air exchange. The model conservatively assumes *k* = 100 as the infection constant (Augenbraun et al., 2020). A higher value may be appropriate for more infectious SARS-CoV-2 strains (Johns Hopkins Medicine, 2021).

It was determined that in-person classes can be safely held in large lecture halls, over the levels of infection rate included in this analysis. However, doing so is not feasible in small classrooms. Although infection probabilities in small classrooms are <1%, the limiting factor is the social distancing requirement. In large lecture halls, this limits student numbers but permits enough students to maintain effective learning environments. Most small classrooms, however, would have insufficient seating capacities following social distancing. This analysis is only valid if universities enforce regulations regarding face mask filtration efficiency, seating capacities, air exchange rates, social distancing, lengths of classes, wait times between classes held in each space, and total number of classes attended in each space over the semester. As evidenced by calculations for Harvard-Cambridge, Harvard-Allston, and Stanford lecture halls, infection risks and required wait times vary by space and should be calculated on a case-by-case basis. These guidelines must also be adapted as infection positivity rates evolve.

In-person dining is largely unsafe, but meal pick-ups can be safely accommodated. The high risk for in-person dining is largely attributable to face mask removal while eating and the significant amount of time involved in this activity. The risk is likely further increased compared to the calculations performed here because the act of eating may increase emissions compared to normal breathing (Papineni & Rosenthal, 1997). Dining halls typically have insufficient air exchange to mitigate this risk. In contrast, meal pick-ups can occur safely if universities enforce regulations regarding face mask filtration efficiency, social distancing, number of students in each meal pick-up batch, length of each meal pick-up window, and wait times between meal pick-up windows. Dining practices observed at universities offering meal-picks are consistent with these findings.

Living in a dorm room with a roommate is highly unsafe if one roommate is infected. Because roommates are in close proximity, and mask wearing is unlikely in personal living spaces and impractical while sleeping, the infection probability is extremely high if one roommate is infected. These findings are consistent with the Harvard College Spring 2021 Plan, which established a maximum of one student per bedroom (Harvard Faculty of Arts and Sciences, 2020).

Bathrooms in university dormitories can be safely shared if universities enforce the conditions used in the modeling of this study. The conclusion of a 40-minute waiting time to minimize infection between uses is consistent, in order of magnitude, with previous analyses, which suggested a 20-minute waiting time (Augenbraun et. al, 2020). The discrepancy is likely related to differing assumptions regarding air exchange rates, mask efficacies, and presence of showers in bathrooms.

The web application developed here allows users to determine infection risk upon inputting several parameters and may provide significant advantage over current risk-assessment methods. Tools such as the Mathematica 19 and Me COVID-19 Risk Calculator (Mathematica, 2020), only provide risk scores based on demographic factors and basic behavioral practices. Other approaches (Augenbraun et al., 2020; Khan et al., 2020) use a variety of infection models and focus on various space types. However, the tool described here is targeted for universities and provides a measure of risk for any teaching space.

## CONCLUSION

With this research, the goal was to quantify SARS-CoV-2 infection risk in various university settings. To that end, a quantitative framework with clear and reasonable assumptions has been created. By creating a web-based tool for users to quantitatively understand the risk they face by inputting the specifications of the space, the model is more accessible to the general public. This tool will continue to be relevant after widespread vaccination because an appreciable risk still exists; vaccines are not 100% effective and some individuals may abstain from vaccination. Given the infection constant and surveillance testing information, this model can be easily adapted to other viruses since the infection constant is the only SARS-CoV-2-specific parameter.

## Data Availability

The code and data used to perform the calculations described in this manuscript are available in Github at https://github.com/mythriambatipudi/RiskAnalysis. Fields marked in gray on the datasheet, however, are private third-party data that cannot be distributed. Access to this data may be granted by the contacts listed in the datasheet.

## RECOMMENDATIONS

The model and web application that have been developed can be utilized to assess the safety of university spaces with regards to SARS-CoV-2 infection risk. Doing so will enable an assessment of the risk of infection for students, faculty, and staff in these spaces and will inform the development and reshaping of university regulations as the COVID-19 pandemic continues to evolve. This model and tool can be used to determine the necessary conditions for safely reopening campuses in order to prevent viral transmission across campuses and maintain safe environments for students, faculty, and staff. Application of this model to specific university campuses and spaces revealed that it is feasible to safely accommodate in-person classes in large lecture halls, meal pick-ups from dining halls and sharing of bathrooms in residential dormitories among small groups of students. However, these conclusions are only true if specific university regulations are enforced. It is especially essential that regulations regarding face mask usage, room capacities, times spent in spaces, wait times between uses of the same space, and air exchange rates are enforced. While several other parameters may also influence the risk of infection, these variables, in particular, must be controlled in order to sufficiently mitigate the risk of transmission. Utilization of the model and web application that have been developed will enable the identification of the exact combination of these parameters that impact the safety of individuals present in each space. While the model and web application have been specifically applied to university spaces, they can also be readily adapted and deployed for the purpose of assessing the level of risk and methods of mitigating this risk in any indoor settings.

## ACKNOWLEDGEMENTS

We would like to acknowledge the Harvard Active Learning Labs and the Harvard Face Mask Committee for providing excellent guidance throughout our investigation process. We would especially like to acknowledge committee members Stephen Blacklow, John Doyle, Willy Shih, Mary Corrigan, Sarah Fortune, and Sara Malconian for their invaluable support and advice. We would also like to recognize Alena Blaise, DaLoria Boone, Jose Gonzalez, Evan Hunsicker, Julia Luehr, Katia Osei, Meghan Turner and James Young for their contributions during the Harvard SEAS engineering design course. Last but not least, we would like to acknowledge the Harvard CS50 course staff for their assistance with publishing the website.

The authors have declared no financial support. The authors have declared no conflicts of interest.

## Notes

### Competing Interest Statement

The authors have declared no competing interest.

### Funding Statement

No funding to report.

## REFERENCES

1. Augenbraun BL, Lasner ZD, Mitra D, Prabhu S, Raval S, Sawaoka H, et al. 2020. Assessment and mitigation of aerosol airborne SARS-CoV-2 transmission in laboratory and office environments. J Occup Environ Hyg. 17(10):447–456. DOI: 10.1080/15459624.2020.1805117.

2. Bar-On YM, Flamholz A, Phillips R, Milo R. 2020. SARS-CoV-2 (COVID-19) by the numbers. Elife. 9:e57309. DOI: 10.7554/eLife.57309.

3. Centers for Disease Control and Prevention, National Center for Immunization and Respiratory Diseases. 2021a. Ventilation in Buildings. Washington (DC): Department of Health and Human Services; [accessed 2021 Jan 8]. https://www.cdc.gov/coronavirus/2019-ncov/community/ventilation.html

4. Centers for Disease Control and Prevention, National Center for Immunization and Respiratory Diseases. 2021b. Your Guide to Masks. Washington (DC): Department of Health and Human Services; [accessed 2021 Jan 8]. https://www.cdc.gov/coronavirus/2019-ncov/prevent-getting-sick/about-face-coverings.html

5. Centers for Disease Control and Prevention, National Institute for Occupational Safety and Health. 2020. Respirator Fact Sheet. Washington (DC): Department of Health and Human Services; [accessed 2021 Jan 8]. https://www.cdc.gov/niosh/npptl/topics/respirators/factsheets/respsars.html

6. United States Environmental Protection Agency. 2020. Ventilation and Coronavirus (COVID-19). Washington (DC): United States Environmental Protection Agency; [accessed 2021 Jan 8]. https://www.epa.gov/coronavirus/ventilation-and-coronavirus-covid-19

7. Fischer EP, Fischer MC, Grass D, Henrion I, Warren WS, Westman E. 2020. Low-cost measurement of face mask efficacy for filtering expelled droplets during speech. Sci Adv. 6(36). DOI: 10.1126/sciadv.abd3083.

8. Hao W, Parasch A, Williams S, Li J, Ma H, Burken J, et al. 2020. Filtration performances of non-medical materials as candidates for manufacturing facemasks and respirators. Int J Hyg Environ Health. 229:113582. DOI: 10.1016/j.ijheh.2020.113582.

9. Harvard Faculty of Arts and Sciences. 2020. Harvard College Spring 2021 Plan. Massachusetts: Harvard University Faculty of Arts and Sciences; [accessed 2021 Jan 9]. https://www.fas.harvard.edu/news/harvard-college-spring-2021-plan.

10. Jayaweera M, Perera H, Gunawardana B, Manatunge J. 2020. Transmission of COVID-19 virus by droplets and aerosols: A critical review on the unresolved dichotomy. Environ Res. 188:109819. DOI: 10.1016/j.envres.2020.109819.

11. Khan K, Bush JW, Bazant MZ. 2020. COVID-19 Indoor Safety Guideline. [accessed 2021 Jan 15]. https://indoor-covid-safety.herokuapp.com/.

12. Konda A, Prakash A, Moss GA, Schmoldt M, Grant GD, Guha, S. 2020. Aerosol Filtration Efficiency of Common Fabrics Used in Respiratory Cloth Masks. ACS Nano. 14(5):6339–6347. DOI: 10.1021/acsnano.0c03252.

13. Mathematica. 2021. 19 and Me: COVID-19 Risk Calculator. New Jersey: Mathematica; [accessed 2021 Jan 8]. https://19andme.covid19.mathematica.org/.

14. Mittal R, Ni R, Seo J-H. 2020. The flow physics of COVID-19. J Fluid Mech. 894. DOI: 10.1017/jfm.2020.330.

15. Papineni RS, Rosenthal FS. 1997. The Size Distribution of Droplets in the Exhaled Breath of Healthy Human Subjects. J Aerosol Med. 10(2):105–116. DOI: 10.1089/jam.1997.10.105.

16. Smalley, A. 2020. Higher Education Responses to Coronavirus (COVID-19). Washington (DC): National Congress of State Legislatures; [accessed 11 Jan 2021]. https://www.ncsl.org/research/education/higher-education-responses-to-coronavirus-covid-19.aspx.

17. The Royal Society. 2020. Face masks and coverings for the general public: Behavioural knowledge, effectiveness of cloth coverings and public messaging. London, UK: The Royal Society; [accessed 24 Sept 2020]. https://royalsociety.org/-/media/policy/projects/set-c/set-c-facemasks.pdf

18. van Doremalen N, Bushmaker T, Morris DH, Holbrook MG, Gamble A, Williamson B, et al. 2020. Aerosol and Surface Stability of SARS-CoV-2 as Compared with SARS-CoV-1. N Engl J Med, 382(16):1564–1567. DOI: 10.1056/NEJMc2004973.

19. Watanabe T, Bartrand TA, Weir MH, Omura T, Haas CN. 2010. Development of a dose-response model for SARS coronavirus. Risk Anal. 30(7):1129–1138. DOI: 10.1111/j.1539-6924.2010.01427.x.

20. Yuen KS, Ye ZW, Fung SY, Chan CP, Jin DY. 2020. SARS-CoV-2 and COVID-19: The most important research questions. Cell Biosci. 10(40). DOI: 10.1186/s13578-020-00404-4.

