## Supplemental Information for "Risk quantification for SARS-CoV-2 infection through airborne transmission in university settings"

**S1 Table. Descriptions of five generic university spaces**

| | Space 1 | Space 2 | Space 3 | Space 4 | Space 5 |
|---|---|---|---|---|---|
| **Positivity Rate** | 0.5% | 1% | 0.7% | 0.9% | 0.6% |
| **Room Volume** | 125,000 $ft^3$ | 20,000 $ft^3$ | 30,000 $ft^3$ | 200,000 $ft^3$ | 20,000 $ft^3$ |
| **Air Exchange Rate** | 1 exchange/hr | 1.5 exchanges/hr | 3 exchanges/hr | 1 exchange/hr | 3 exchanges/hr |
| **Filtration Efficiency of Face Mask Worn** | 80% | 70% | 90% | 60% | 75% |
| **Normal Room Capacity** | 500 | 100 | 20 | 1000 | 5 |
| **Time per Visit** | 1.5 hr | 1 hr | 1.5 hr | 1.5 hr | 5 min |
| **Total # of Visits per Semester** | 180 | 252 | 180 | 180 | 420 |

** Note if one of these space variables is the x variable for a graph in Figures 3 and 4, the variable will cover the full range of values indicated in the graph rather than the value listed in the table here.*
